# Age-dependent increase in tau phosphorylation at serine 396 in Huntington’s disease pre-frontal cortex

**DOI:** 10.1101/2023.06.03.23290851

**Authors:** Tiziana Petrozziello, Sommer S. Huntress, Ayleen L. Castillo-Torres, James P. Quinn, Theresa R. Connors, Corinne A. Auger, Alexandra N. Mills, Spencer E. Kim, Sophia Liu, Farah Mahmood, Adel Boudi, Muzhou Wu, Ellen Sapp, Pia Kivisäkk, Shekar R. Sunderesh, Mahmoud A. Pouladi, Steven E. Arnold, Bradley T. Hyman, H. Diana Rosas, Marian DiFiglia, Ricardo Mouro Pinto, Kimberly Kegel-Gleason, Ghazaleh Sadri-Vakili

**Affiliations:** Sean M. Healey &AMG Center for ALS at Mass General, Mass General Brigham, Boston, MA 02129, USA; Department of Neurology, Mass General Brigham, Boston, MA 02129, USA; Center for Genomic Medicine, Massachusetts General Brigham, Boston, MA 02114, USA; Department of Medical Genetics, Centre for Molecular Medicine and Therapeutics, British Columbia Children’s Hospital Research Institute, University of British Columbia, Vancouver, V5Z 4H4, Canada; Department of Neurology, Harvard Medical School, Boston, MA 02115, USA

**Keywords:** Huntington’s disease, total tau, phosphorylated tau, fractionations, post-mortem brain, ESC-derived neurons, NSCs, mouse models, plasma

## Abstract

**Background:** To date, it is still controversial whether tau phosphorylation plays a role in Huntington’s disease (HD), as previous studies demonstrated either no alterations or increases in phosphorylated tau (pTau) in HD post-mortem brain and mouse models.

**Objectives:** The goal of this study was to determine whether total tau and pTau levels are altered in HD.

**Methods:** Immunohistochemistry, cellular fractionations, and western blots were used to measure tau and pTau levels in a large cohort of HD and control post-mortem prefrontal cortex (PFC). Furthermore, western blots were performed to assess tau, and pTau levels in HD and control isogenic embryonic stem cell (ESC)-derived cortical neurons and neuronal stem cells (NSCs). Similarly, western blots were used to assess tau and pTau in *Htt^Q111^* and transgenic R6/2 mice. Lastly, total tau levels were assessed in HD and healthy control plasma using Quanterix Simoa assay.

**Results:** Our results revealed that, while there was no difference in tau or pTau levels in HD PFC compared to controls, tau phosphorylated at S396 levels were increased in PFC samples from HD patients 60 years or older at time of death. Additionally, tau and pTau levels were not changed in HD ESC-derived cortical neurons and NSCs. Similarly, tau or pTau levels were not altered in *Htt^Q111^* and transgenic R6/2 mice compared to wild-type littermates. Lastly, tau levels were not changed in plasma from a small cohort of HD patients compared to controls.

**Conclusion:** Together these findings demonstrate that pTau-S396 levels increase significantly with age in HD PFC.

## Introduction

Huntington’s disease (HD) is an autosomal dominant neurodegenerative disease caused by a cytosine-adenosine-guanine (CAG) trinucleotide expansion coding for a polyglutamine (polyQ) repeat in the huntingtin (Htt) protein [1]. The disease is characterized by severe motor impairment together with or followed by cognitive and psychiatric symptoms [1, 2]. Although the genetic cause of HD is well-characterized, the pathogenic mechanisms leading to neuronal loss are not yet fully elucidated.

Recent studies have suggested that the microtubule-associated protein (MAP) tau may be involved in HD pathogenesis [4–6]. Tau plays a critical role in a group of neurodegenerative diseases known as tauopathies, such as Alzheimer’s disease (AD), progressive supranuclear palsy and some frontotemporal lobe degenerations [7, 8]. In these disorders, mutations in *MAPT*, the gene encoding tau, or post-translational modifications of the tau protein cause abnormal accumulation of tau in both neurons and glia, disrupting crucial cellular functions [8–10]. For example, phosphorylation of key tau epitopes including S396/S404 and S202/T205, cause the formation of neuropil thread and neurofibrillary tangles (NFTs) [8, 10] that disrupt neuronal transport, an early pathogenetic event in neurodegenerative diseases [6, 7, 11], including HD [12–14].

Although genome-wide association studies (GWAS) have not revealed HD-specific mutations or polymorphisms in *MAPT* [15], studies assessing total tau and phosphorylated tau (pTau) levels as well as the presence of NFTs in HD have demonstrated conflicting results. Increases in tau and tau phosphorylated at both the microtubule binding sites and the proline-rich domains have been reported in HD [3–5]. Furthermore, while region-specific alterations in pTau and NFTs were previously described in post-mortem HD brains [4–5, 14, 16–20], NFTs were not observed in the caudate, temporal and frontal lobes in post-mortem HD samples in a study by Singhrao and colleagues [21].

Similarly, inconsistent findings have been reported on tau levels in cerebrospinal fluid (CSF) from people living with HD. In one study increases in CSF tau were reported in HD [22, 23], while in others CSF tau levels were reported not changed; however, there was a positive correlation between tau and mutant huntingtin (Htt) levels in CSF [24, 25]. Therefore, it is still unclear whether tau could serve as a viable biomarker in HD.

In light of the contradictory results on the role of tau in HD, here we examined potential changes in tau and its phosphorylated form at S396 (pTau-S396), S404 (pTau-S404), and T181 (pTau-T181) in a large cohort of human post-mortem PFC, a brain region that is also affected in HD and extensively studied by our group. Furthermore, isogenic embryonic stem cell (ESC)-derived cortical neurons and neuronal stem cells (NSCs) from people living with HD and controls were assessed for changes in tau and pTau. Total tau and pTau levels were also assessed in the cortex of a full length, *Htt^Q111^*, and fragment R6/2 mouse models of HD. Lastly, we measured tau levels in a small cohort of HD plasma samples and compared them to healthy sex- and age-matched controls to assess the role of tau as a potential biomarker for HD.

## Materials and Methods

### Human brain tissue samples

Post-mortem PFC from HD and controls were obtained from the Massachusetts Alzheimer’s Disease Research Center (ADRC) with approval from the Mass General Brigham Institutional Review Board (IRB). In total we assessed 38 HD, and 29 control PFC. The mean age was 80.27 years (SD = 11.54) for the control group and 57.51 years (SD = 11.84) for the HD group. Post-mortem interval (PMI) range was 8-72 h for the control group and 4-24 h for the HD group.

### Immunohistochemistry and Image Analysis

Seven-µm-thick paraffin-embedded brain sections were immunostained for pTau-S396 (1:100; #ab109390, Abcam, MA) using a Bond Rx autostainer (Leica Biosystems, IL), according to the manufacturer’s instructions and as previously reported [26]. Briefly, slides were batch processed with the following settings: Bake and Dewax, immunohistochemistry (IHC) protocol F 60 minutes, HIER 20 minutes with ER1. Slides were then transferred into water and dehydrated by 1-minute incubations into baths of 70% ethanol, 95% ethanol, 100% ethanol, and xylene. Slides were then cover slipped using Permount Mounting Medium (Fisher Scientific) and left to dry overnight. Slides were scanned using an NanoZoomer Digital Pathology-HT scanner (C9600-12; Hamamatsu Photonics, Shizuoka, Japan) at a magnification of 20X. Scanned slide images were visualized in NDP.view2 viewing software (Hamamatsu, Shizuoka, Japan) and analyzed in ImageJ 1.53a (National Institute of Health, Bethesda, MD).

### Western blotting

Western blots were performed using previously described protocols [26–28]. Briefly, 50 μg of proteins were resuspended in sample buffer and separated on a 4-12% bis-tris protein gel for 90 min at 120V. Proteins were then transferred to a PVDF membrane in an iBlot Dry Blotting System (Invitrogen, Thermo Fisher, MA), and the membrane was blocked with 5% bovine serum albumin (BSA) in tris-buffered saline with Tween 20 (TBST) before immunodetection with the following primary antibodies: total tau (1:1000; #A0024, DAKO, Denmark), pTau-S396 (1:500; #ab109390, Abcam, MA), PSD-95 (1:500; #2507S, Cell Signaling, MA), and GADPH (1:1000; #MAB0009, Millipore Sigma, Termecula, CA) overnight at 4°C. Primary antibody incubation was followed by 4 washes in TBST before incubation with the secondary antibody for 1 h (HRP-conjugated goat anti-rabbit IgG, and HRP-conjugated goat anti-mouse IgG; Jackson ImmunoResearch Laboratories, West Grove, PA). After 4 washes in TBST, proteins were visualized using the ECL detection system (Thermo Fisher Scientific, MA). Protein levels were then analyzed using Image J 1.53a (National Institute of Health, Bethesda, MD).

### Sarkosyl fractionation

Fractionation was performed using an adapted protocol from Matsumoto et al., 2015 [29]. Human post-mortem PFC (370 mg) were homogenized in 3.7 mL homogenization buffer (100mg/mL) composed of 10 mM Tris-HCl, 0.8 M NaCl, 1 mM EGTA, and 1 mM dithiothreitol (DTT) supplemented with protease and phosphate inhibitors cocktail. Seven hundred μL were saved as total homogenate fraction, while the remaining samples were then centrifuged at 100,000 g for 20 min at RT. The supernatants were collected and saved as total soluble fraction, while the pellets were resuspended and sonicated in homogenization buffer containing 1% Triton X-100 and incubated at 37^0^ C for 30 min. At the end of the incubation, the samples were centrifuged at 100,000 g for 20 min at RT. While the supernatants were collected and saved as Triton X-100 soluble fraction, the pellets were resuspended and sonicated in 500 μL of homogenization buffer containing 1% Sarkosyl and incubated at 37^0^ C for 30 min. Next, the samples were centrifuged again at 100,000 g for 20 min and the supernatants were collected and saved as Sarkosyl-soluble fraction. The pellets instead were resuspended and sonicated in 320 μL of 20 mM Tris-HCl buffer, pH 7.5. These samples were lastly collected and saved as Sarkosyl-insoluble fraction. Bradford assay was used to determine protein concentration in total, total soluble, Triton X-100 soluble, Sarkosyl soluble, and Sarkosyl insoluble fractions. Tau and pTau-S396 levels were detected by western blots and stained with amido black as a loading control.

### Synaptoneurosomes (SNs) fractionation

SNs fractionation was performed as previously reported [30, 31]. Human post-mortem PFC (200 to 300mg) were homogenized in 500 μL ice-cold Buffer A composed of 25 mM Hepes pH 7.5, 120 mM NaCl, 5 mM KCl, 1mM MgCl_2_, 2 mM CaCl_2_, 2 mM DTT, 1 mM NaF supplemented with phosphate and protease inhibitors cocktail. The homogenates were then passed through two layers of 80μm nylon filters (Millipore, MA) to remove tissue debris. Seventy μL aliquot was saved, mixed with 70 μL H_2_O and 23 μL 10% sodium dodecyl sulfate (SDS), passed through a 27-gauge needle to shear DNA, boiled for 5 min, and centrifuged at 15,000g for 15 min to prepare the total extract. The remainder of the homogenates was passed through a 5μm Supor membrane Filter (Pall Corp, Port Washington, NY) to remove organelles and nuclei, centrifuged at 1000g for 5 min and both pellet and supernatant were saved. The supernatants were collected in small crystal centrifuge tubes and centrifuged at 100,000g for 45 min at 4^0^C to obtain the cytosolic fractions. The pellets were resuspended in Buffer B composed of 50 mM Tris pH 7.5, 1.5% SDS and 2 mM DTT, boiled for 5 min, and centrifuged at 15,000 g for 15 min to obtain SNs. Bradford assay was used to determine protein concentration in total extract, cytosolic fraction, and SNs.

### Embryonic stem cells, neural stem cell and cortical neuron differentiations

Oversight of all stem cell work occurred by the Human Embryonic Stem Cell Research Oversight Committee (ESCRO Committee) and by the MassGeneral Brigham (MGB) Institutional Biosafety Committee (PIBC) (ESCRO #2015-01-02 and PIBC Reg #2017B000023). The isogenic human embryonic stem cells (ESCs) H9 (WA09, WiCell) have CAG repeats in the HTT alleles of 19/30 and were used to engineer new lines using Talen that contain 19/65 CAGs [32]. The method created two base pair changes in the coding region of HTT which are silent (do not result in a change at the amino acid level). The karyotype of these cells was normal by array comparative genomic hybridization (aCGH, Agilent 60K Standard, Cell Line Genetics, WI). Human ESCs were maintained in mTeSR™ Plus kit (#100-0276, StemCell Technologies, Vancouver, Canada) on Matrigel (#354277, Corning, MA). Differentiation to neural stem cells (NSCs) was carried out as previously described [33]. ESCs were differentiated into cerebral cortex neurons using previously published protocols [34] with few modifications. Briefly, neural induction occurred on confluent EPS plated on Matrigel (#354277, Corning, MA) for 10 days in Neural Induction Medium (NIM) Neuroepithelial sheets were lifted with dispase (#07923, StemCell Technologies, Vancouver, Canada) and plated on Matrigel. FGF treatment (20 ng/mL; #PHG0023, Gibco, ThermoFisher Scientific, MA) in Neural Maintenance Medium (NMM) was initiated on day 14 and stopped at Day 18 where neural rosettes were observed.

Upon successful neural development, cells were cultured continuously in NMM with BDNF (10ng/mL; #PHC7074, Gibco, ThermoFisher Scientific, MA) and GDNF (10 ng/mL, #PHC7075, Gibco, ThermoFisher Scientific, MA) 42 days after the start of the neural induction along with anti-mitotic inhibitors 5-Fluoro-2’-Deoxyuridine (1 µM; #F0503, Sigma-Aldrich, MO) and Udirine (1 µM, #U3750, Sigma-Aldrich, MO).

### Animals

All animal work was performed according to the guidelines set by the Massachusetts General Hospital Subcommittee on Research Animal Care (SRAC).

#### Htt^Q111^ mice

*Htt^Q111^* knock in (KI) mice contain a humanized *Htt* exon 1with a sequence encoding an expanded polyQ tract (∼111 glutamines) as well as the human polyproline tract [35, 36]. Six-month-old wild-type and homozygous (*Htt^Q111^*) mice were sacrificed, and brains were rapidly removed and dissected to isolate the cortex. Cortices were then frozen in liquid isopentane and stored at -80°C until use.

#### R6/2 mice

R6/2 transgenic mice contain exon 1 of the HD gene with an expanded CAG repeat (150-200) under control of the human HD promoter [37]. Twelve-week-old R6/2 transgenic and wild-type littermate controls were sacrificed, and brains were rapidly removed and dissected to isolate the cortex. Cortices were then frozen in liquid isopentane and stored at -80°C until use.

### Human Plasma samples

Plasma samples from healthy control participants were obtained from the LifeSPAN biorepository at the Mass General Institute for Neurodegenerative Disease (MIND). Plasma samples from HD participants were collected by venipuncture using EDTA containing tubes. Samples were spun in a refrigerated centrifuge at 1000 g; plasma was removed and respun at 25,000 g in order to remove any remaining red blood cells. The supernatant was carefully removed, aliquoted into low protein binding cryovials and frozen at -80C until processed. In total, we assessed plasma samples from 10 participants living with HD and 10 age- and gender-matched healthy controls.

### Quanterix Simoa assays

Plasma total tau concentration was measured in duplicate using the Simoa Tau Advantage Kit on a fully automated Simoa HD-X Analyzer using manufacturer’s instructions (Quanterix Corporation, Billerica, MA) and as previously reported [26] in the MADRC Biomarkers core. Duplicate coefficient of variations (CVs) were 5.6±4.3% (mean±SD), and all samples had a CV<15%. One sample had concentration under functional lower level of quantification (LLOQ) of 0.38 pg/mL.

### Statistics

Normal distribution of data was not assumed regardless of sample size or variance. Individual value plots with the central line representing the median and the whiskers representing the interquartile range, and box plot with the central line representing the median, the edges representing the interquartile range, and the whiskers representing the minimum and maximum values, were used for graphical representation. Comparisons between groups were performed using a non-parametric Mann-Whitney U test, and a two-way ANOVA followed by Tukey’s test. Correlation of CAG repeat length with tau, pTau-S396, pTau-S404 and pTau-T181 levels were performed as non-parametric Spearman correlations. All tests were two sided with a significance level of 0.05, and exact p values are reported. Analyses were performed using GraphPad Prism 9.0.

### Study approval

The study was approved by the Partners Healthcare Institutional Review Board and Institutional Animal Care and Use Committee. Written informed consent was obtained from all participants prior to study enrollment. Post-mortem consent was obtained from the appropriate representative (next of kin or health care proxy) prior to autopsy.

## Results

### pTau-S396 levels were increased in elderly HD PFC

In order to determine whether there were alterations in tau phosphorylation in HD, we first assessed the levels of tau, pTau-S396, pTau-S404, and pTau-T181 in HD and control PFC by western blots. Our results revealed no differences in total tau, pTau-S396, pTau-S404, and pTau-T181 levels between HD and control PFC (Figure 1).

**Figure 1.**
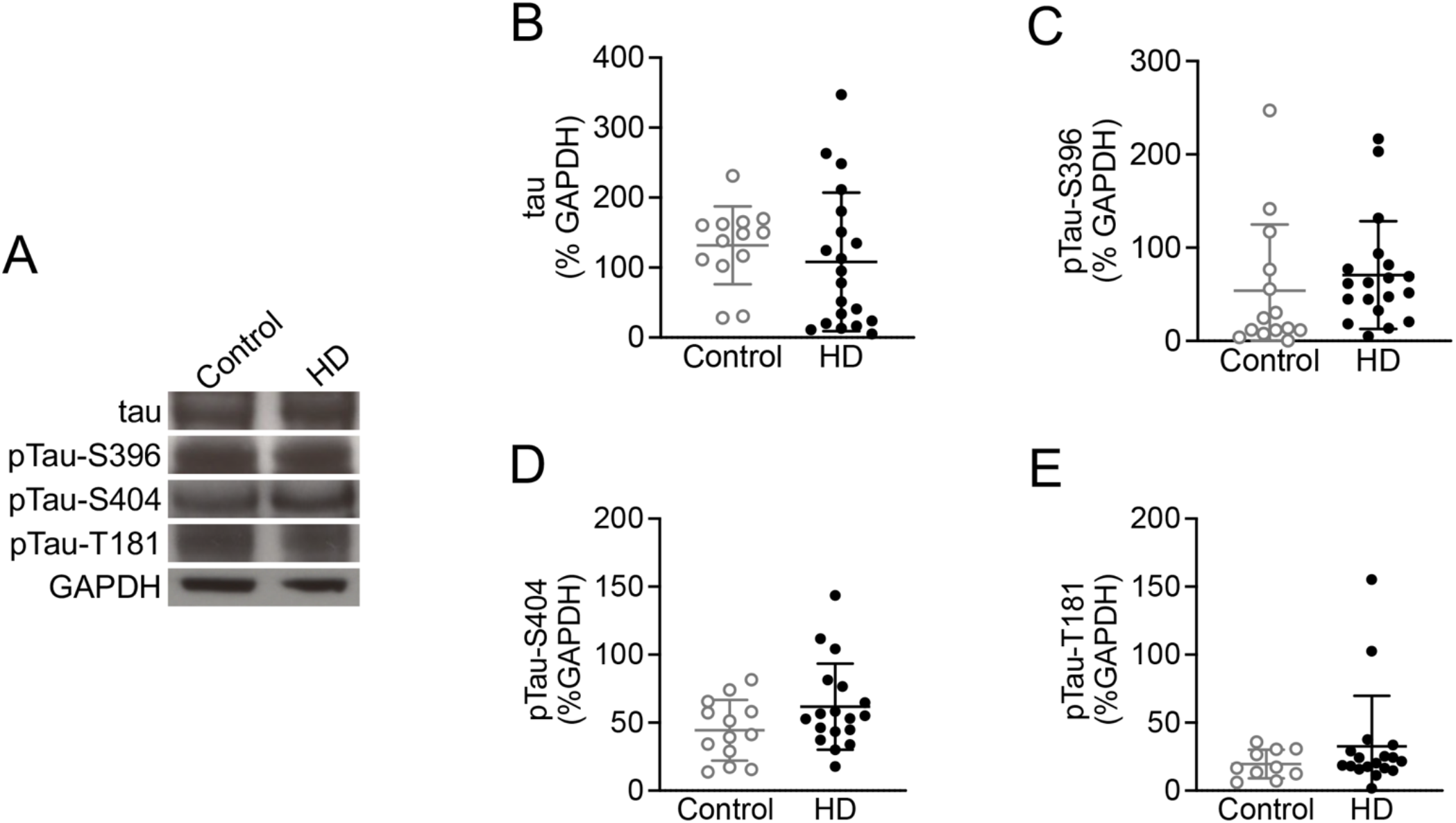
Total tau and pTau-S396 levels were not altered in HD PFC. (**A**) Representative western blot images of tau, pTau-S396, pTau-S404, pTau-T181 and GAPDH in whole cell homogenates derived from control and HD PFC revealing no alterations in tau and pTau levels. (**B**) There were no alterations in tau in HD PFC (n = 20) compared to controls (n = 14) (Mann-Whitney U test = 95, p = 0.2069). (**C**) pTau-S396 levels were not altered in HD PFC (n = 20) compared to controls (n = 14) (Mann-Whitney U test = 87.50, p = 0.0997). (**D**) There were no alterations in pTau-S404 between HD (n = 19) and control PFC (n = 14) (Mann-Whitney U test = 82, p = 0.1697). (**E**) pTau-T181 levels were not altered in HD PFC (n = 19) compared to controls (n = 11) (Mann-Whitney U test = 71.50, p = 0.3879).

Next, to determine whether there was an effect of CAG length on tau and pTau levels in HD, we evaluated the association between total tau, pTau-S396, pTau-S404, and pTau-T181 levels with CAG repeat length for each sample. Our results revealed no impact of the CAG repeat length on either tau, pTau-S396, pTau-S404, or pTau-T181 levels in PFC (Suppl. Figure 1).

Given that previously published studies demonstrate that tau phosphorylation was increased in elderly HD patients [3, 15, 18, 19, 38, 39], we next subdivided our samples based on age at the time of death, young (< 60 y) and elderly (> 60 y). Our results revealed that while there were no differences in pTau-S396 levels in PFC derived from younger patients, there was a significant increase in pTau-S396 levels in PFC derived from elderly HD patients compared to age-matched controls (Figure 2A and B). However, there were no changes in pTau-S404 and pTau-T181 in either young or elderly HD PFC compared to age-matched control PFC (Figure 2C-F).

**Figure 2.**
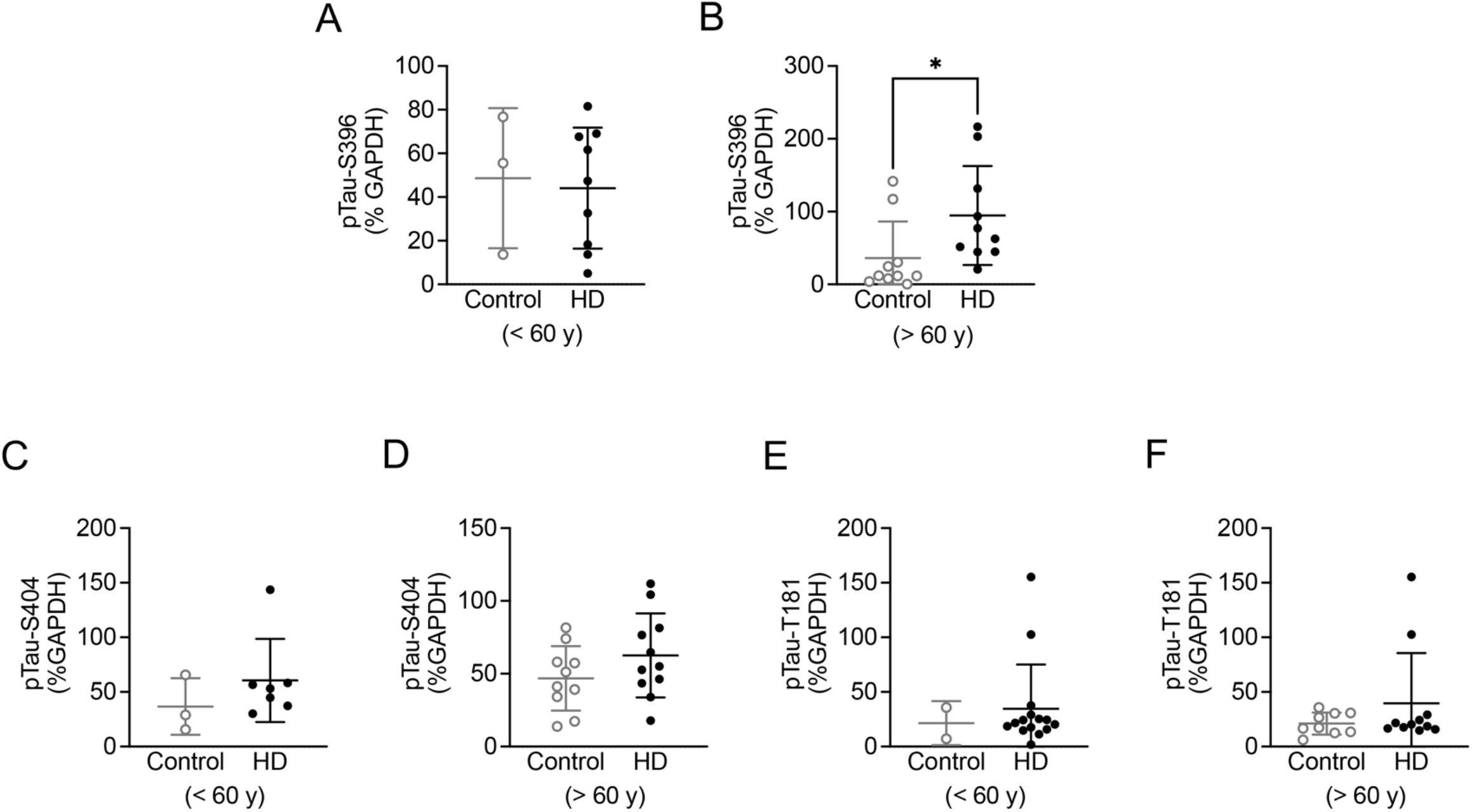
pTau-S396 levels were increased in elderly HD PFC. (**A**) pTau-S396 levels did not change in PFC derived from younger HD patients (< 60 y; n = 9) compared to age-matched controls (n = 3) (Mann-Whitney U test = 12.50, p = 0.8955). (**B**) There was a significant increase in pTau-S396 levels in PFC derived from older HD patients (> 60 y; n = 11) compared to age-matched controls (n = 10) (Mann-Whitney U test = 17, p = 0.0110). (**C**) No alterations in pTau-S404 levels were found in young HD PFC (n = 7) compared to controls (n = 3) (Mann-Whitney U test = 6, p = 0.3833). (**D**) pTau-S404 levels were not altered in older HD PFC (n = 12) compared to age-matched controls (n = 11) (Mann-Whitney U test = 38, p = 0.2512). (**E**) There was no change in pTau-T181 levels between HD (n = 7) and control PFC (n = 2) derived from younger patients (Mann-Whitney U test = 6, p = 0.8889). (**F**) pTau-T181 levels were not altered in older HD PFC (n = 12) compared to controls (n = 9) (Mann-Whitney U test = 44.50, p = 0.7250). * p < 0.05.

To further investigate and confirm our findings, we focused on pTau-S396 and assessed its levels by IHC in a smaller young (< 60 y) cohort of HD post-mortem PFC based on tissue availability. An entorhinal cortex sample from an AD brain was used as positive control. As expected, extensive neuropil threads and NFTs were observed in the AD cortex, while no threads or NFTs were evident in HD or control PFC (Figure 3A and C). Furthermore, the analysis revealed no significant difference in pTau-S396 levels between HD and control PFC in both grey and white matter (Figure 3B and D). Lastly, no impact of the CAG repeat size on pTau-S396 levels was found in either grey or white matter (Suppl. Figure 2).

**Figure 3.**
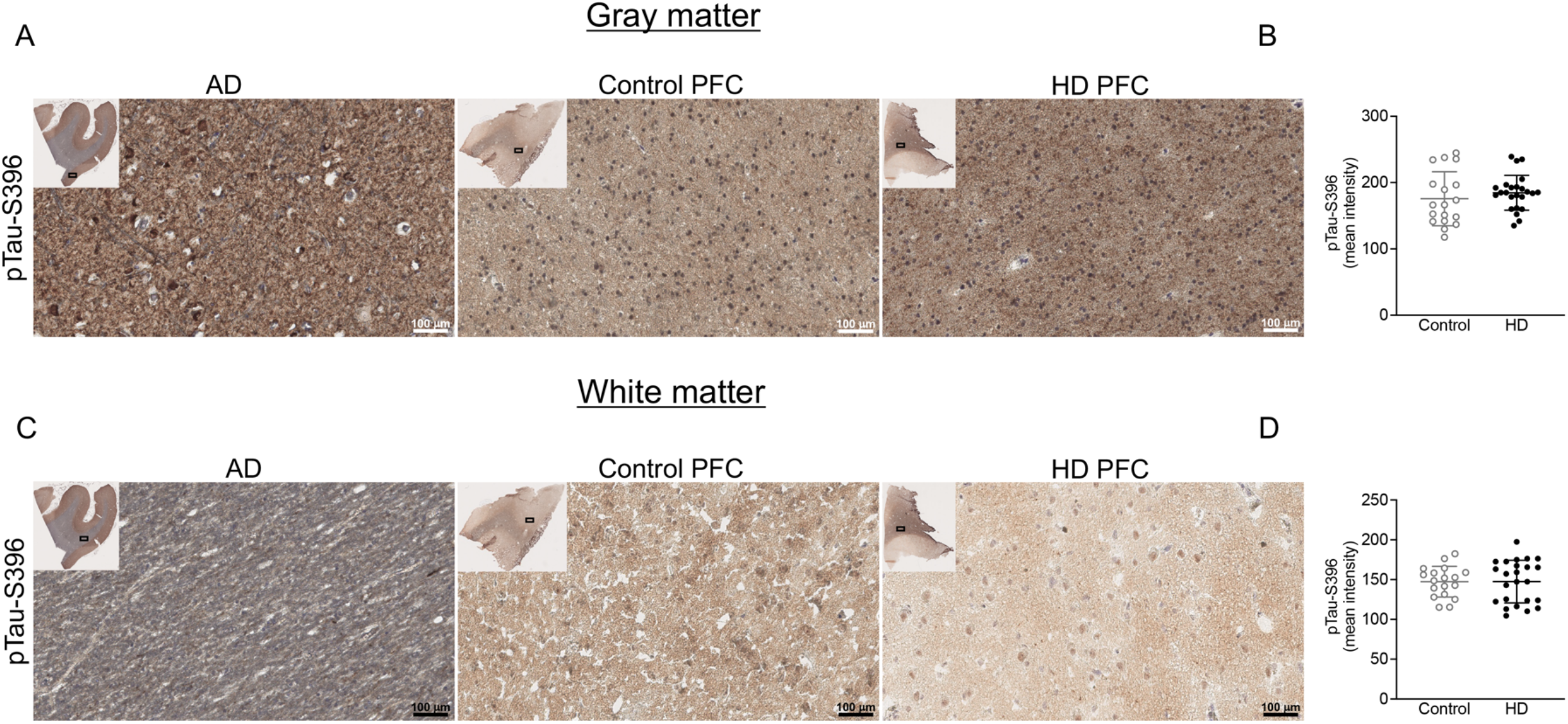
pTau-S396 levels were not altered in gray and white matter in HD PFC. (**A**) Representative IHC images of pTau-S396 staining in grey matter from AD entorhinal cortex, control and HD PFC. Extensive neuropil threads and NFTs were observed in AD cortex, while no threads or NFTs were evident in both control and HD PFC. (**B**) There were no alterations in pTau-S396 levels in the grey matter from HD PFC (n = 24) compared to controls (n = 18) (Mann-Whitney U test = 175, p = 0.3069). (**C**) Representative IHC images of pTau-S396 staining in the white matter from AD entorhinal cortex, control and HD PFC. Neuropil threads and NFTs were not observed in control and HD PFC. (**D**) No significant changes were observed in pTau-S396 levels in HD PFC white matter (n = 24) compared to control PFC (n = 18) (Mann-Whitney U test = 210.5, p = 0.8950). Scale bar: 100 μm; insert scale bar: 5 mm.

### Total tau and pTau-S396 levels were not altered in soluble and insoluble fractions in HD post-mortem PFC

To assess the levels of soluble and insoluble species of tau and pTau, we next performed a Sarkosyl fractionation, and measured tau and pTau-S396 levels by western blots in a smaller young (< 60 y) cohort of HD post-mortem PFC based on tissue availability. The results demonstrated that there was no significant change in tau and pTau-S396 levels in total extract, total soluble, Triton X-100 soluble, and the Sarkosyl soluble fractions in HD compared to control PFC (Figure 4A-L). Furthermore, tau and pTau-S396 levels were not altered in the Sarkosyl insoluble fractions in HD PFC compared to controls (Figure 4M-O). Similarly, there was no effect of the CAG repeat length on either tau or pTau-S396 levels in any of the fractions (Suppl. Figure 3).

**Figure 4.**
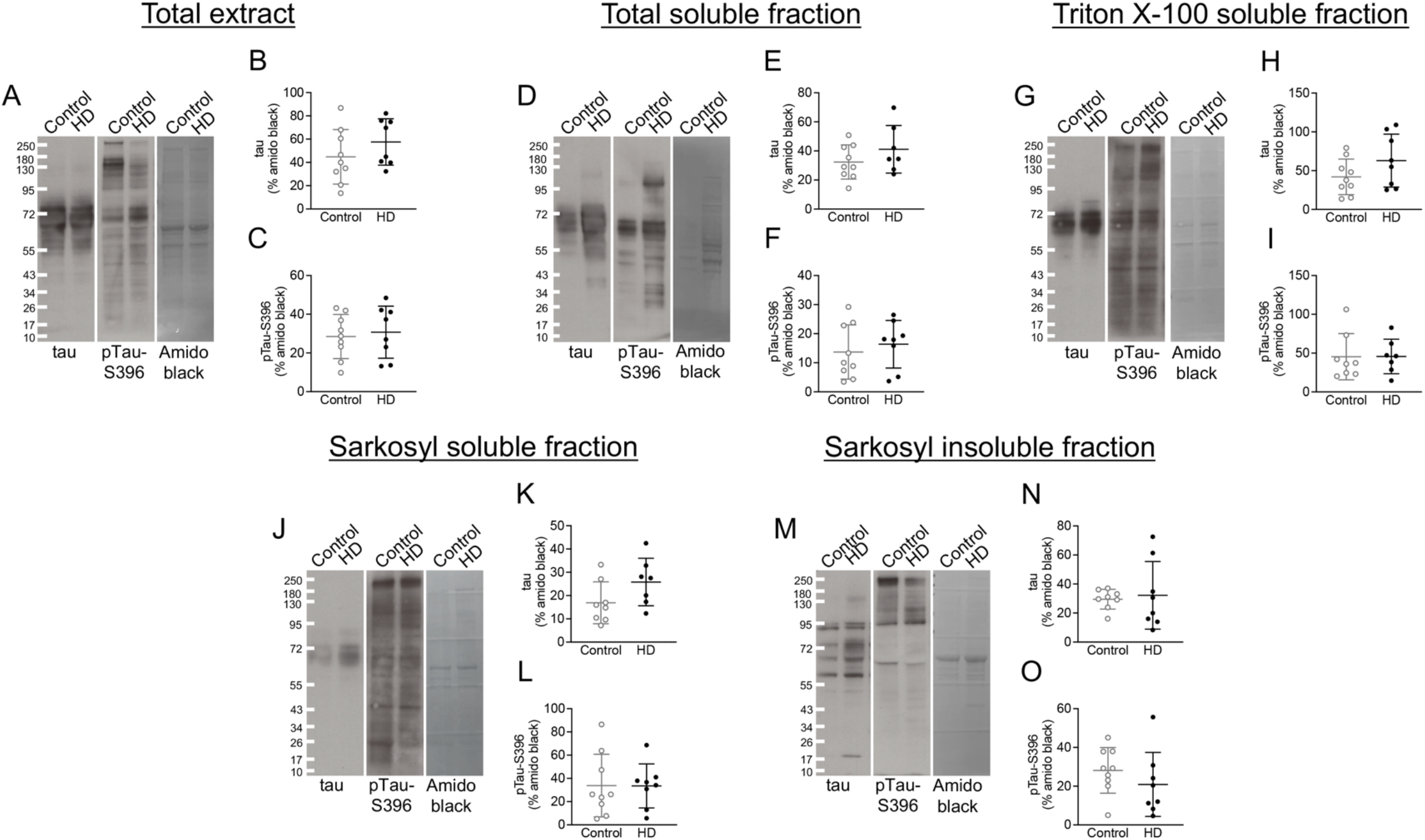
Soluble and insoluble species of total tau and pTau-S396 were not altered in HD PFC. (**A**) Representative western blot images of tau, pTau-S396 and amido black in total extract fractions derived from control and HD PFC revealing no alterations in tau and pTau-S396. (**B**) No differences were observed in tau levels in the total extract fraction from HD PFC (n = 8) compared to controls (n = 9) (Mann-Whitney U test = 23, p = 0.2359). (**C**) There were no changes in pTau-S396 levels in total extract fraction from HD (n = 8) and control PFC (n = 9) (Mann-Whitney U test = 33, p = 0.8148). (**D**) Representative western blot images of tau, pTau-S396 and amido black in total soluble fractions from control and HD PFC demonstrating no changes in tau and pTau-S396. (**E**) Tau levels were not changed in HD total soluble fraction (n = 8) compared to controls (n = 9) (Mann-Whitney U test = 24, p = 0.4698). (**F**) No significant alterations were observed in pTau-S396 levels in HD total soluble fraction (n = 8) compared to control soluble fraction (n = 9) (Mann-Whitney U test = 29, p = 0.5414). (**G**) Representative western blot images of tau, pTau-S396 and amido black in Triton X-100 soluble fractions derived from control and HD PFC revealing no alterations in tau and pTau-S396. (**H**) There were no changes in tau levels in the Triton X-100 soluble fraction from HD PFC (n = 8) and controls (n = 9) (Mann-Whitney U test = 23, p = 0.2359). (**I**) No alterations were found in pTau-S396 levels in Triton X-100 soluble fraction between HD PFC (n = 8) and controls (n = 9) (Mann-Whitney U test = 23, p = 0.6126). (**J**) Representative western blot images of tau, pTau-S396 and amido black in Sarkosyl soluble fractions from control and HD PFC demonstrating no differences in tau and pTau-S396. (**K**) Tau levels were not altered in the Sarkosyl soluble fractions in HD (n = 8) compared to control PFC (n = 9) (Mann-Whitney U test = 12, p = 0.0721). (**L**) There were no changes in pTau-S396 levels in the Sarkosyl soluble fraction in HD (n = 8) compared to control PFC (n = 9) (Mann-Whitney U test = 31, p = 0.6730). (**M**) Representative western blot images of tau, pTau-S396 and amido black in Sarkosyl insoluble fractions from control and HD PFC revealing no alterations in tau and pTau-S396. (**N**) Tau levels were not changed in the Sarkosyl insoluble fractions derived from HD (n = 8) and control PFC (n = 9) (Mann-Whitney U test = 27, p = 0.6454). (**O**) There was no change in pTau-S396 levels in the Sarkosyl insoluble fraction between HD PFC (n = 8) and controls (n = 9) (Mann-Whitney U test = 23, p = 0.2359).

### Total tau and pTau-S396 levels were not altered at the synaptic level in HD PFC

Given the increase in synaptic pTau in AD and amyotrophic lateral sclerosis (ALS) [30, 31], we next assessed the levels of tau and pTau-S396 in three different PFC fractions – total extract, cytosol, and SNs – from the younger cohort of HD PFC by western blots. The purity of the fractionation was demonstrated by the detection of PSD-95 in the total and SNs fractions, while no PSD-95 was detected in the cytosolic fraction derived from both control and HD PFC (Figure 5A). Our results revealed no significant difference in the levels of total tau and pTau-S396 in any of the fractions between HD and control PFC (Figure 5B and C). Additionally, no impact of the CAG repeat length on either tau or pTau-S396 levels was observed in the synaptic fractions (Suppl. Figure 4).

**Figure 5.**
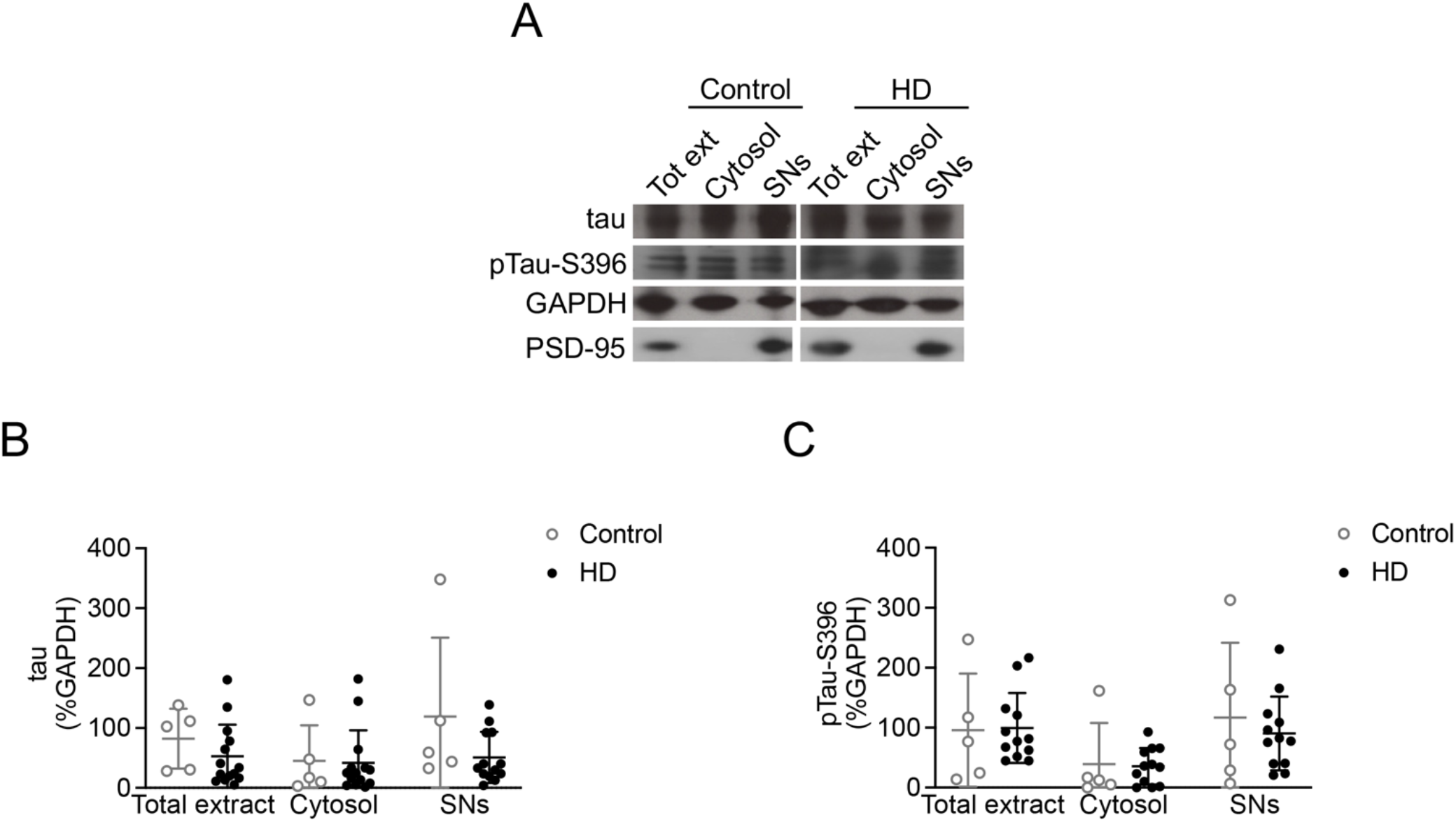
Synaptic total tau and pTau-S396 levels were not altered in HD PFC. (**A**) Representative western blot images of tau, pTau-S396, PSD-95, and GAPDH in total extracts, cytosolic fractions and SNs derived from control and HD PFC. There were no alterations in tau and pTau-S396 levels between HD and control PFC. PSD-95 immunosignal was detected in total extracts and SNs demonstrating purity of the fractionation. (**B**) Two-way ANOVA revealed no significant effect of cellular fractions ([F(2,58) = 1.812], p = 0.1724), genotype [F(1,58) = 0.2429], p = 0.6240) and cellular fraction X genotype interaction ([F(2,58) = 0.2701], p = 0.7643) on tau levels. Tukey’s test confirmed no change in tau levels in the three fractions derived from HD (n = 14) and control PFC (n = 5) (p > 0.05). (**C**) Two-way ANOVA demonstrated a significant effect of cellular fractions ([F(2,45) = 4.133], p = 0.0255), while there were no significant effects of genotype [F(2,45) = 0.1766], p = 0.6763) and cellular fraction X genotype interaction ([F(2,45) = 0.1895], p = 0.8280) on pTau-S396 levels. Tukey’s test revealed no alterations between the three fractions in HD (n = 15) and control samples (n = 5) (p > 0.05).

### Total tau and pTau-S396 levels are not altered in HD ESC-derived cortical neurons or NSCs

We next measured total tau and pTau levels in cortical neuron cultures and NSCs derived from isogenic control or HD ESCs. The isogenic pair expressed Htt with a polyQ expansion in the normal range (21/17Q) or with one expanded repeat (65Q/17Q). Our results revealed no significant difference in total tau and pTau-S396 levels in HD ESC-derived cortical neuronal cultures compared to the isogenic control line (Figure 6A-C).

**Figure 6.**
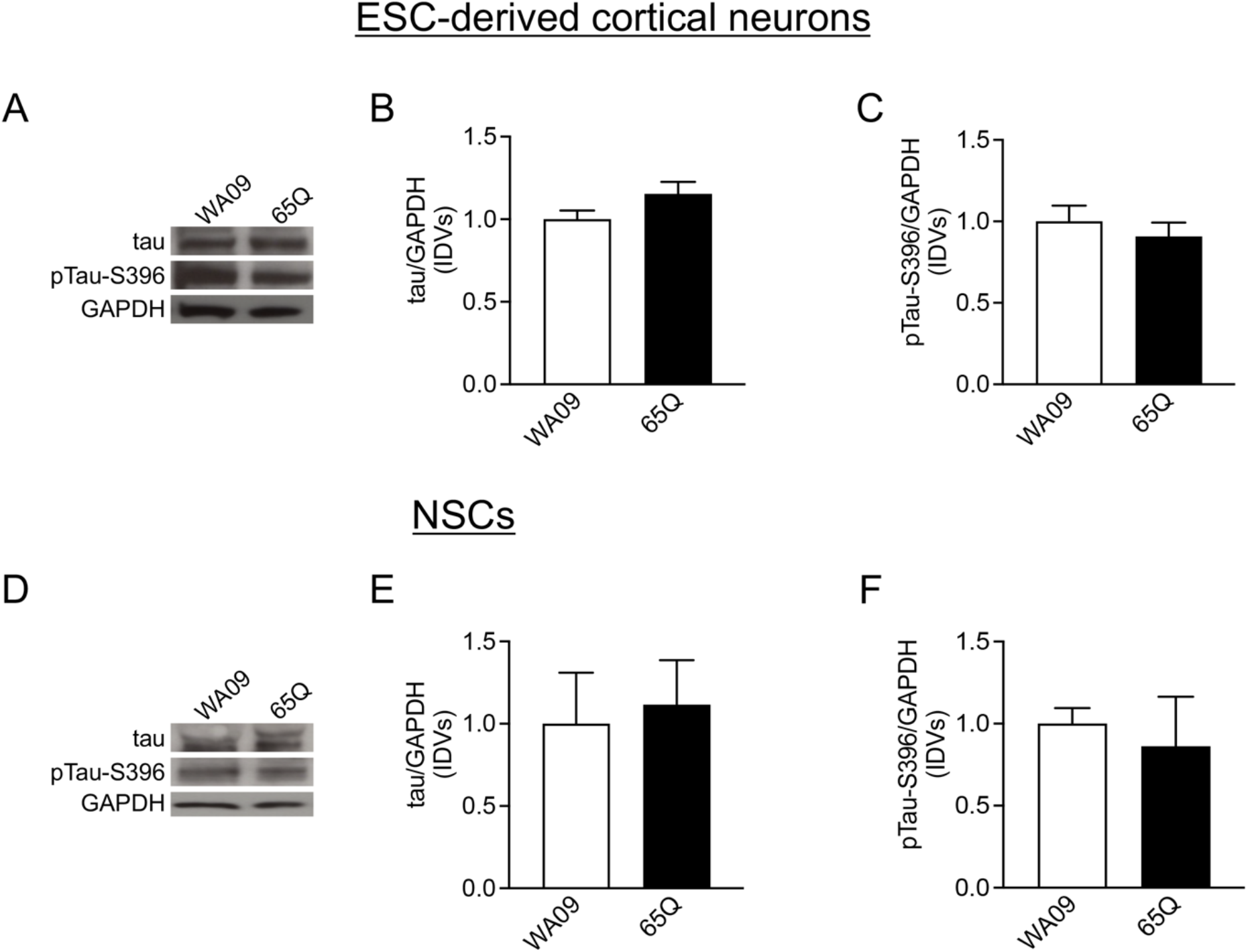
There were no alterations in total tau and pTau-S396 levels in HD ESC-derived cortical neurons and NSCs. (**A**) Representative western blot images of tau, pTau-S396, and GAPDH in ESC-derived cortical neurons from HD and controls revealing no alterations in tau and pTau-S396 levels. (**B**) There were no significant differences in total tau levels in HD ESC-derived neurons compared to the isogenic control cell line (Mann-Whitney U test = 2, p = 0.1143). (**C**) No differences in pTau-S396 levels were observed in ESC-derived cortical neurons between HD and control cell lines (Mann-Whitney U test = 6, p = 0.6857). (**D**) Representative western blot images of tau, pTau-S396, and GAPDH in NSCs derived from HD and control patients demonstrating no differences in tau and pTau-S396 levels. (**E**) There were no alterations in total tau levels in HD ESC-derived NSCs compared to isogenic control cells (Mann-Whitney U test = 4, p >0.9999). (**F**) pTau-S396 levels did not change in HD NSCs compared to isogenic control cells (Mann-Whitney U test = 3, p = 0.7000).

Similarly, no significant differences were reported in the levels of both pTau-S404 and pTau-T181 in HD ESC-derived cortical neurons compared to the isogenic control cell line (Suppl. Figure 5A-C). Additionally, there were no significant alterations in total tau and pTau-S396 levels in NSCs derived from HD and isogenic control cells (Figure 6D-F). Similarly, there were no alterations in pTau-S404 and pTau-T181 levels in HD NSCs compared to isogenic control NSCs (Suppl. Figure 5D-F).

### There were no differences in total tau and pTau-S396 levels in two HD mouse models

Total tau and pTau levels were measured by western blots in cortical samples derived from *Htt^Q111^* knock-in and R6/2 mice. The results revealed that there was no difference in total tau or pTau-S396 levels between *Htt^Q111^* compared to wild-type mice (Figure 7A-C) as well as transgenic R6/2 mice compared to wild-type littermates (Figure 7D-F). Similarly, there was no change in the levels of pTau-S404 and pTau-T181 between *Htt^Q111^*and transgenic R6/2 mice compared to their wild-type littermate (Suppl. Figure 6).

**Figure 7.**
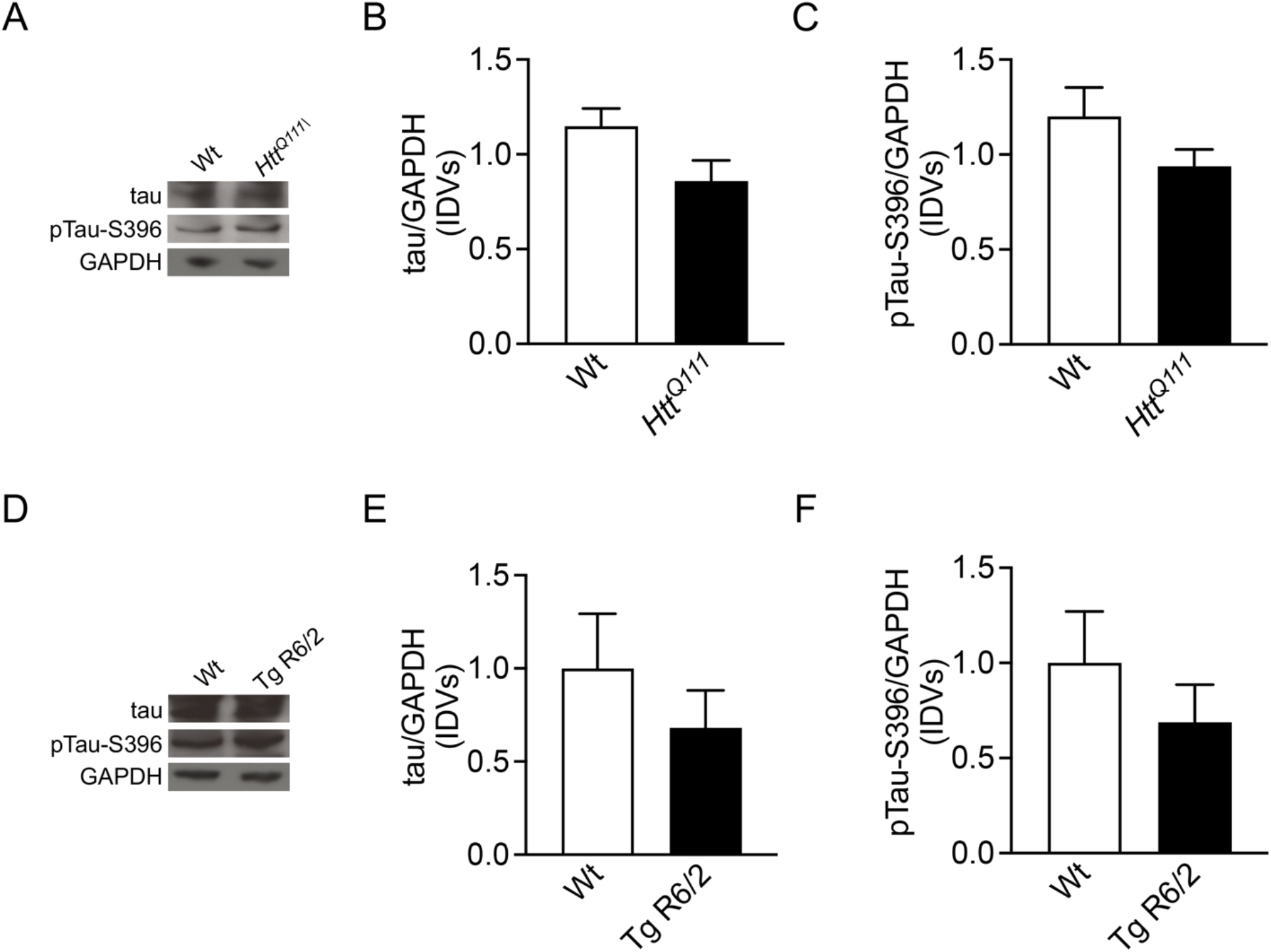
There were no differences in total tau and pTau-S396 levels in two different HD mouse models. (**A**) Representative western blot images of tau, pTau-S396 and GAPDH in wild-type (Wt) and *Htt^Q111^* cortex revealing no alterations in tau and pTau-S396 levels. (**B**) No changes in tau levels were reported in *Htt^Q111^* (n = 8) compared to Wt cortex (n = 8) (Mann-Whitney U test = 15, p = 0.0830) (**C**) There were no alterations in pTau-S396 levels in *Htt^Q111^* (n = 8) compared to Wt cortex (n = 8) (Mann-Whitney U test = 18, p = 0.1521). (**D**) Representative western blot images of tau, pTau-S396 and GAPDH in R6/2 transgenic mice and Wt littermates demonstrating no differences in tau and pTau-S396 levels. (**E**) No alterations in total tau levels were observed in transgenic R6/2 (Tg) (n = 8) compared to Wt cortex (n = 8) (Mann-Whitney U test = 28, p = 0.7209). (**F**) pTau-S396 levels were not altered in Tg R6/2 (n = 8) compared to Wt cortex (n = 8) (Mann-Whitney U test = 23, p = 0.3671).

### Plasma tau levels were not altered in HD

Plasma tau has been suggested as a viable biomarker of disease (Lee at al., 2019). Therefore, total tau levels were measured in plasma samples obtained from people living with HD and age- and sex-matched control using Quanterix Simoa assay. The analysis revealed no differences in plasma total tau levels in HD compared to controls (Figure 8).

**Figure 8.**
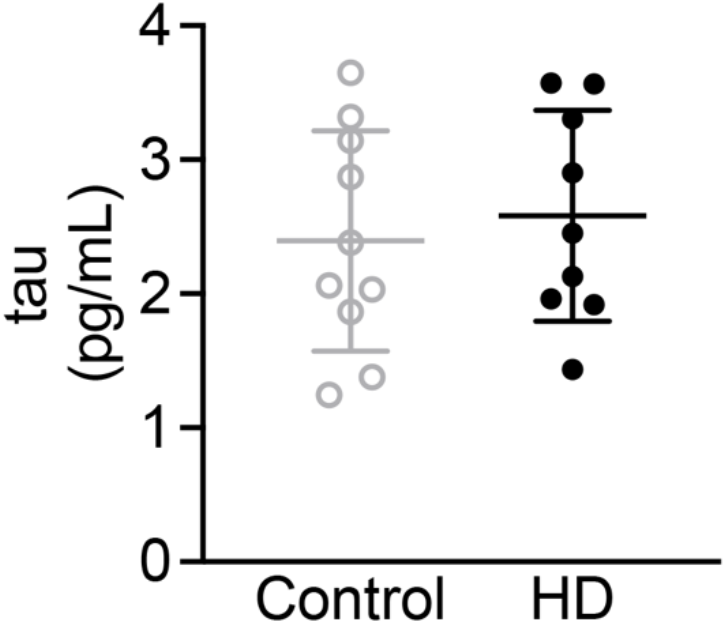
Tau levels were not altered in HD plasma. There were no alterations in plasma tau levels between HD (n = 9) and healthy control patients (n = 10) (Mann-Whitney U test = 38, p = 0.6038). One sample had a tau level under the LLOQ and was not included in the figure.

## Discussion

In this study, we report no changes in total tau and its phosphorylated forms at S396, S404, and T181 in HD post-mortem PFC, human ESC-derived cortical neurons and NSCs as well as two mouse models of HD. Furthermore, we were unable to detect neuropil threads and NFTs in either the grey or white matter in the HD PFC samples by using an antibody against pTau-S396. Similarly, there was no significant change in the levels of soluble and insoluble or synaptic total tau or pTau-S396 in HD PFC. Although our results revealed no association between CAG length and total tau, pTau-S396, pTau-S404 or pTau-T181 levels in HD, pTau-S396 levels in PFC derived from older HD patients (> 60 y) were significantly increased. Lastly, plasma tau levels remain unchanged in HD.

In this study, we did not detect an overall increase in tau and pTau, supporting the previously published results of Singhrao and colleagues revealing no NFTs in caudate, temporal and frontal lobes in HD [21]. However, our results contradict other reports indicating the presence of neuropil threads and NFTs in post-mortem HD brain regions [15, 18–20, 38, 39]. This may be due to differences in study design: first, in this study we only assessed the PFC in post-mortem samples; second, another key difference is the choice of pTau antibodies. In our study, we used antibodies against pTau-S396, pTau-S404 and pTau-T181, while the majority of prior studies demonstrating NFTs in post-mortem HD brains used the AT8 antibody which recognizes phosphorylated tau at S202 and T205. We did try using AT8 and additional pTau antibodies; however, we were unable to detect a signal in our samples. Therefore, future studies are required to assess additional brain regions such as the striatum as well as additional pTau antibodies to further clarify the role of tau phosphorylation in HD.

Increases in pTau have been described in older HD patients and over time during the course of the disease [3, 15, 18, 19, 38, 39], a finding that our study also supports. Indeed, although we did not report an overall increase in the levels of total tau or pTau-S396 in HD PFC, our results demonstrate a significant increase in pTau-S396 levels in older patients (> 60 y) compared to age-matched controls. Furthermore, we did not observe any significant impact of CAG repeat length on either total tau or pTau-S396 levels, further suggesting that increases in pTau may be an age-related event in HD.

In this study, we did not detect any differences in soluble or insoluble total tau or pTau-S396 in HD PFC compared to controls. Additionally, we did not report any impact of the CAG repeat length on total tau or pTau-S396 levels in either soluble or insoluble fractions. Although insoluble tau is the main component of NFTs, it has been reported that both soluble and insoluble species of tau exert toxic effects in the central nervous system [40]. Specifically, in HD, previous studies have demonstrated an increase in insoluble tau species in post-mortem cortex and striatum from HD patients [3, 4] together with a decrease in soluble tau [3]. Although our findings are consistent with our data demonstrating no overall change in tau and pTau-S396 in HD PFC, we did use a smaller cohort of samples for the fractionation experiments. Furthermore, we used a young (< 60 y) cohort of HD PFC for all fractionation studies due to tissue availability. Although the results are consistent and demonstrate that there are no differences in tau or pTau in cellular fractions derived from younger HD PFC, we were not able to determine whether changes in soluble or insoluble species of tau are age-dependent in HD for these studies. Here, we also assessed synaptic tau levels given that our recent finding in ALS demonstrated an accumulation of hyperphosphorylated tau at the synaptic level similar to AD [30, 31]. It has been suggested that this accumulation may contribute to synapse loss and impairments in trafficking [31] which are also early pathogenic events in HD occurring prior to symptom onset (12, 41-45). However, our data did not reveal a significant change in synaptic total tau and pTau-S396 levels in HD PFC. Similarly, there was no effect of the CAG repeat length on either total tau and pTau-S396 levels at the synaptic levels in HD. Similar to the Sarkosyl fractionation, we used a smaller and younger (< 60 y) cohort of HD PFC for the SNs fractionation. Therefore, future studies using older samples are necessary to determine whether synaptic pTau-S396 is increased in elderly HD patients.

We further confirmed our findings using cellular models of HD which demonstrated that total tau and pTau-S396 levels are not altered. Indeed, our results revealed no change in tau and pTau-S396 levels in ESC-derived cortical neurons with an expanded CAG repeat compared to its isogenic control cell line. Similarly, no differences were reported in NSCs derived from the same isogenic pair. However, one caveat of these results is that embryonic neuron cultures are very young and still developing. Therefore, studies in aged neurons are required to validate our findings. Our transgenic mouse studies also did not demonstrate any differences in the levels of total tau and pTau-S396 in the cortex of 12-week-old transgenic R6/2 mice as well as in 11-month-old knock in *Htt^Q111^* mice, contrary to previous studies revealing an increase in tau phosphorylation in 10-week-old R6/2 mice [46, 47]. Collectively, our results in cellular and animal models of HD support and confirm our findings in post-mortem brain.

Lastly, we measured plasma tau in a small cohort of HD and healthy control samples. Tau could provide a viable biomarker for the diagnosis of tauopathies, such as AD, given that increases in tau levels have been described in patient biofluids, including CSF and plasma [48–50]. We also have recently demonstrated that CSF tau levels correlate with faster disease progression in ALS [26], further supporting the role of tau as biomarker for neurodegenerative diseases. However, we did not detect a difference in tau levels in our HD samples; therefore, it is still unclear whether tau may serve as diagnostic or prognostic biomarkers as previously suggested in studies revealing increasing in tau in CSF from people living with HD [22, 23]. Furthermore, in two additional studies, it was shown that while CSF tau levels were not altered in HD, there was a positive correlation between tau and mutant Htt [24, 25]. It is possible, as the first pilot study to measure plasma tau levels in HD, our sample size might have been too small to detect any meaningful differences. Future studies using a larger sample size are necessary to validate our findings in plasma and to determine whether tau may serve as a viable biomarker for HD.

Taken together, our results indicate that there is not an overall increase in tau and pTau in HD post-mortem PFC, HD cellular and animal models as well as in plasma from people living with HD. However, the increase in pTau-S396 in older HD patients supports the idea that increases in tau phosphorylation may be linked to aging in HD.

## Data Availability

All data produced in the present study are available upon reasonable request to the authors.

## Author contributions

T.P. contributed to the study design, data collection, data analysis, and drafting of the manuscript. S.S.H., A.L.C.-T., J.P.Q., T.R.C., C.A.A., A.N.M., S.E.K., S.L., F.M., A.B., M.W., E.S., P.K., S.R.S., M.A.P. contributed to the data collection, data analysis, and editing of the manuscript. S.E.A., B.T.H., H.D.R., M.D., R.M.P., K.K.-G. contributed to the study design and editing of the manuscript. G.S.-V. contributed to the study design, data analysis, drafting of the manuscript.

## Acknowledgments

The authors would like to thank patients and their family for sample donation. M.D., H.D.R., R.M.P., K.K.-G. and G.S-V. were supported by the Dake Family Foundation. M.A.P. was the recipient of a BC Children’s Hospital Research Institute Investigator Grant Award (IGAP), and a Scholar Award from the Michael Smith Health Research BC. The MADRC/MIND Biomarker Core and Biorepository was supported by NIA grant P30AG062421 (S.E.A.). R.M.P. was supported by the National Institute of Neurological Disorders and Stroke, National Institute of Health (NS126420).

## Data availability statement

The data supporting the findings of this study are available within the article and/or its supplementary material.

---

**Supplementary Figure 1.**
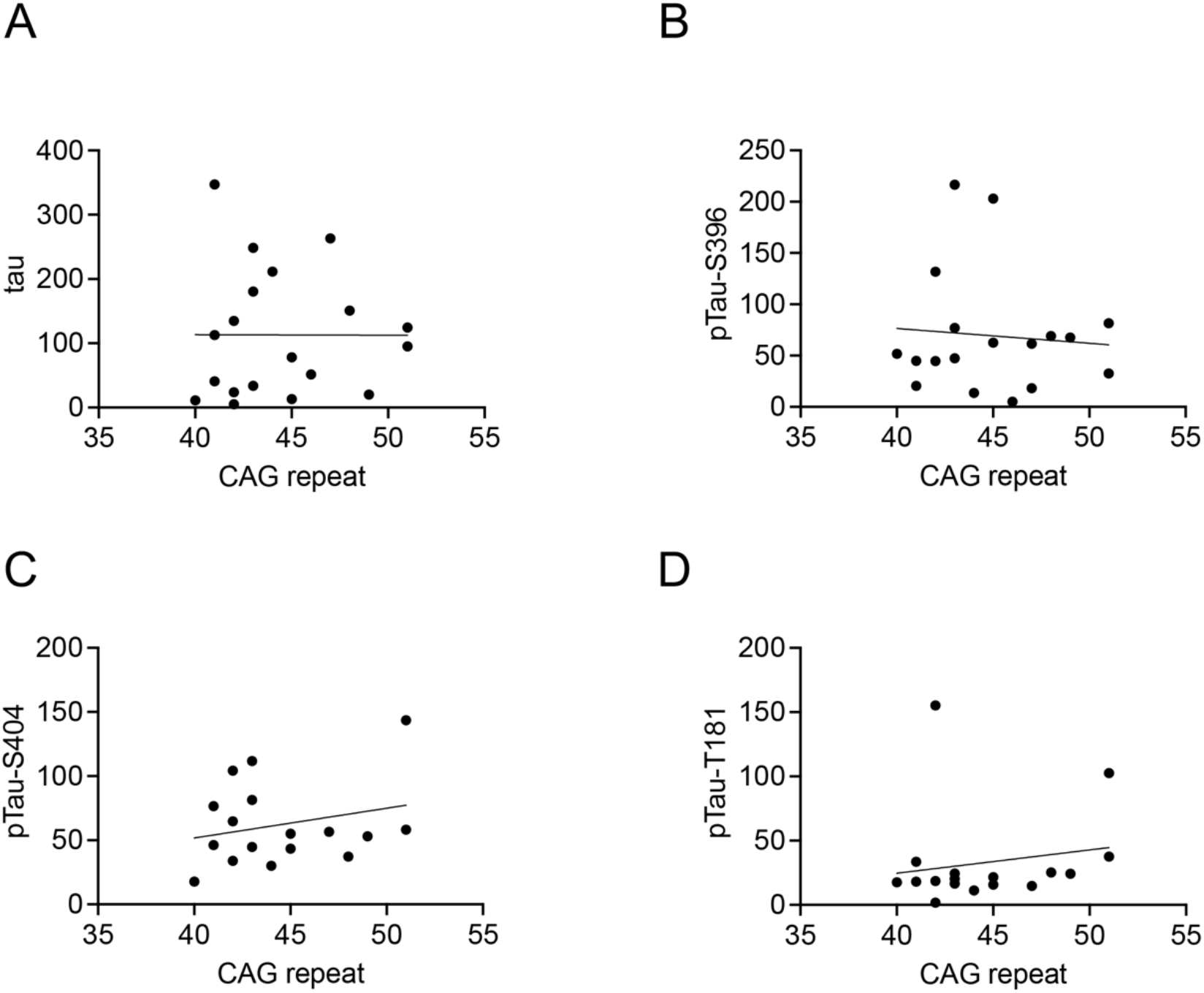
There was no impact of the CAG repeat length on either total tau, pTau-S396, pTau-S404 or pTau-T181 levels in HD PFC. (**A**) There was no effect of the CAG repeat length on total tau levels in HD PFC (n = 20) (Spearman correlation, R = 0.1342, p = 0.5840). (**B**) There was no impact of the CAG repeat length on pTau-S396 levels in HD PFC (n = 20) (Spearman correlation, R = 0.05702, p = 0.8222). (**C**) CAG repeat length had no effect on the levels of pTau-S404 in HD PFC (n = 20) (Spearman correlation, R = 0.1505, p = 0.5615). (**D**) No effect of CAG repeat length was observed on pTau-T181 levels in HD PFC (n = 20) (Spearman correlation, R = 0.2418, p = 0.3471).

**Supplementary Figure 2.**
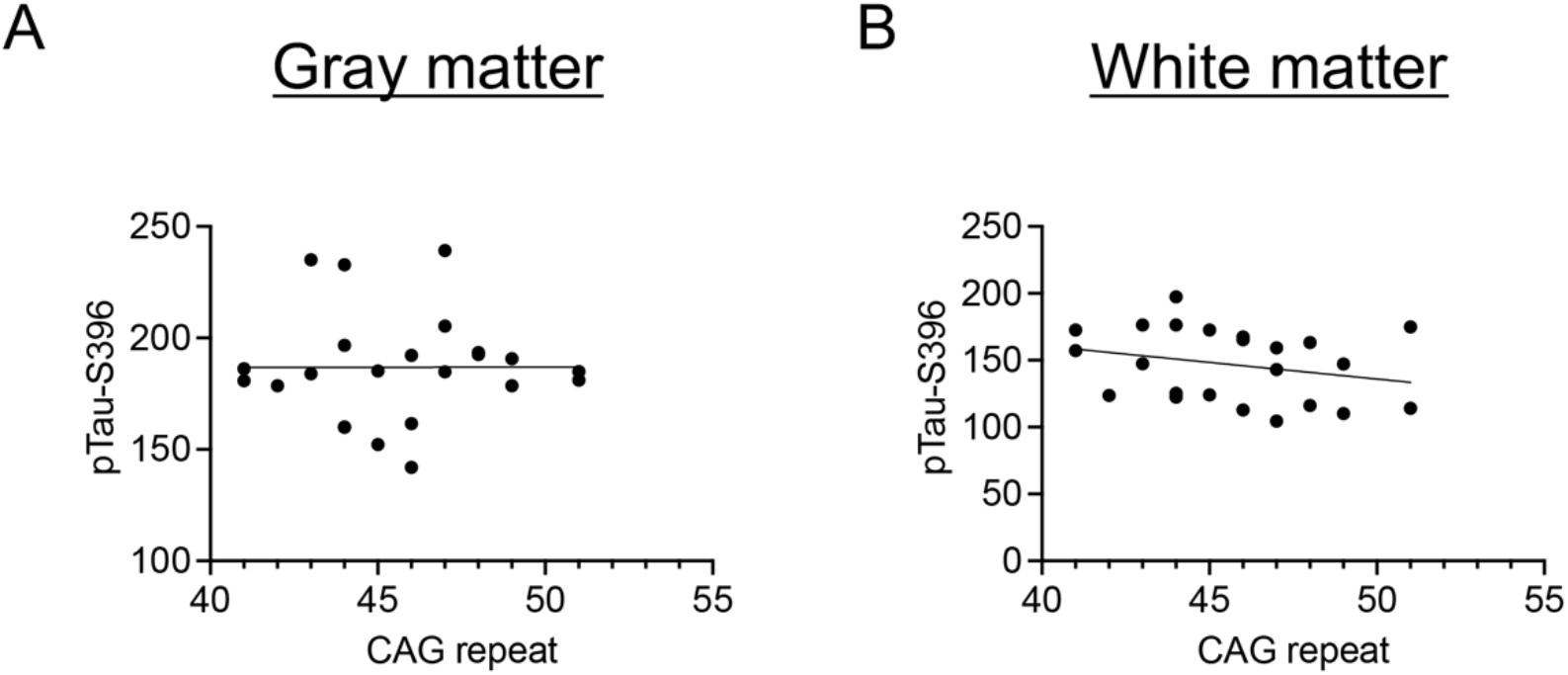
No effect of the CAG repeat length was observed on pTau-S396 levels in grey and white matter from HD PFC. (**A**) There was no impact of the CAG repeat length on the levels of pTau-S396 in HD PFC grey matter (n = 24) (Spearman correlation, R = 0.1133, p = 0.6067). (**B**) There was no effect of the CAG repeat length on pTau-S396 levels in the HD PFC white matter (n = 24) (Spearman correlation, R = -0.3236, p = 0.1319).

**Supplementary Figure 3.**
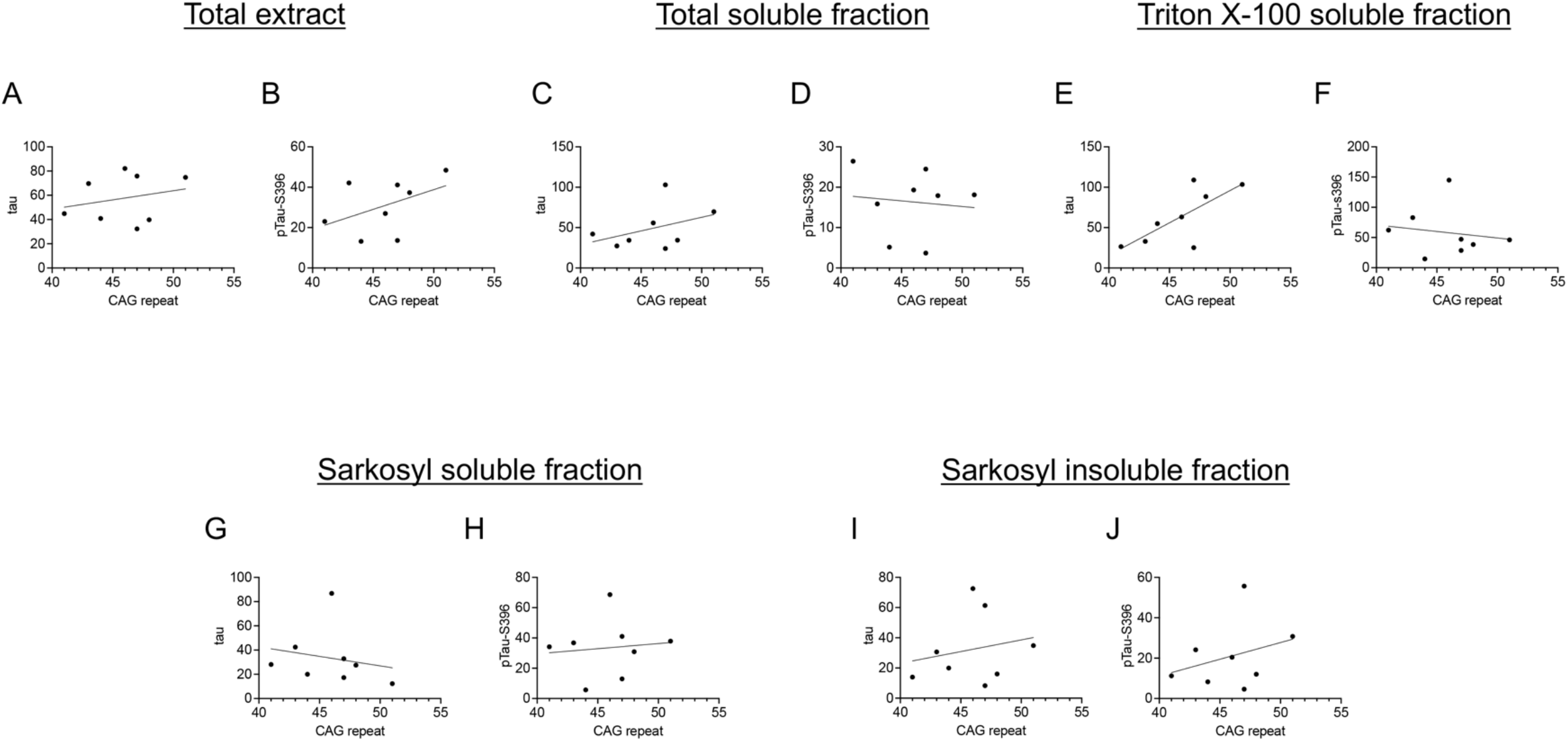
There was no effect of the CAG repeat length on total tau and pTau-S396 levels in soluble and insoluble fractions from HD PFC. (**A**) There was no effect of CAG repeat length on total tau levels in the total extract fraction (n = 8) (Spearman correlation, R = -0.02395, p = 0.9640). (**B**) CAG repeat length had no impact on pTau-S396 levels in total extract fraction (n = 8) (Spearman correlation, R = 0.4072, p = 0.3168). (**C**) There was no effect of the CAG repeat length on the levels of total tau in the total soluble fraction (n = 8) (Spearman correlation, R = 0.3234, p = 0.4344). (**D**) There was no impact of CAG repeat on pTau-S396 levels in the total soluble fraction (n = 8) (Spearman correlation, R = -0.1437, p = 0.7349). (**E**) CAG repeat length did not have any effect on total tau levels in the Triton X-100 soluble fraction (n = 8) (Spearman correlation, R = 0.6108, p = 0.1158). (**F**) There was no impact of CAG repeat on pTau-S396 levels in the Triton X-100 soluble fraction (n = 8) (Spearman correlation, R = -0.3713, p = 0.3651). (**G**) There was no effect of CAG repeat length on the levels of total tau in the Sarkosyl soluble fraction (n = 8) (Spearman correlation, R = -0.5270, p = 0.1847). (**H**) CAG repeat had no effect on pTau-S396 levels in the Sarkosyl soluble fraction (n = 8) (Spearman correlation, R = 0.1317, p = 0.7606). (**I**) CAG repeat length did not have any impact on total tau levels in the Sarkosyl insoluble fraction (n = 8) (Spearman correlation, R = 0.1677, p = 0.6922). (**J**) There was no effect of the CAG repeat on pTau-S396 levels in the Sarkosyl insoluble fraction (n = 8) (Spearman correlation, R = 0.2994, p = 0.4714) in HD PFC.

**Supplementary Figure 4.**
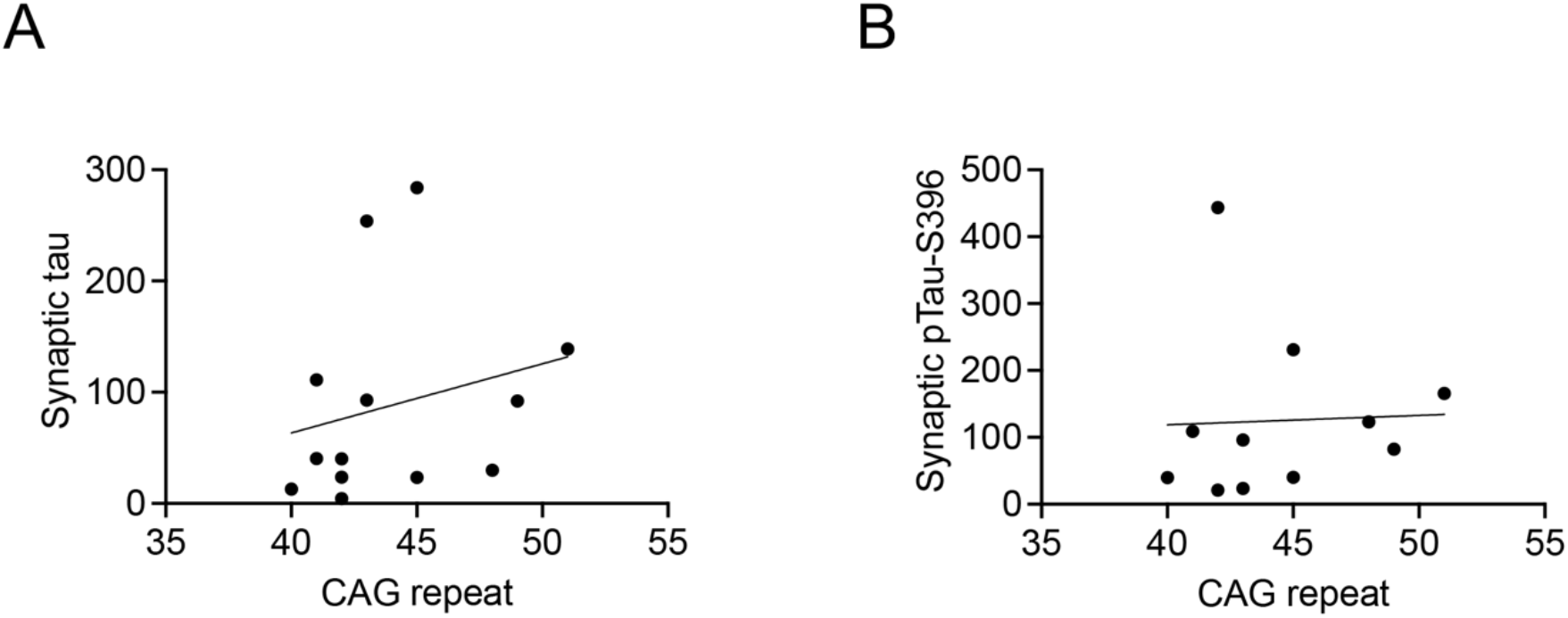
There was no impact of the CAG repeat length on synaptic total tau and pTau-S396 levels in HD PFC. (**A**) There was no effect of the CAG repeat length on synaptic total tau in HD PFC (n = 15) (Spearman correlation, R = 0.3662, p = 0.2175). (**B**) CAG repeat length had no effect on synaptic pTau-S396 levels in HD PFC (n = 15) (Spearman correlation, R = 0.3021, p = 0.3641).

**Supplementary Figure 5.**
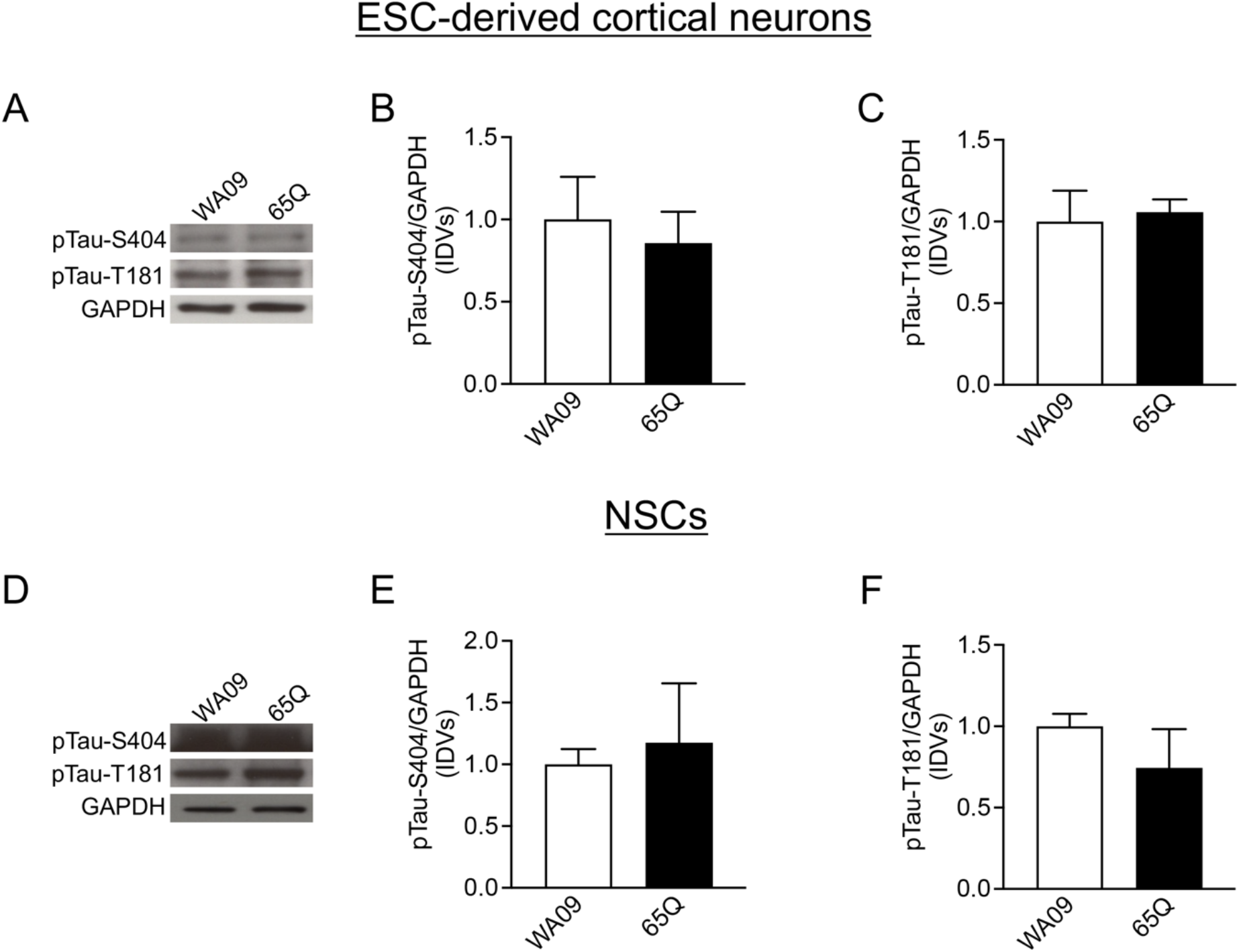
There were no alterations in pTau-S404 and pTau-T181 levels in HD ESC-derived cortical neurons and NSCs. (**A**) Representative western blot images of pTau-S404, pTau-T181, and GAPDH in HD and control ESC-derived cortical neurons revealing no alterations in pTau-S404, and pTau-T181 levels. (**B**) There were no significant differences in pTau-S404 levels in HD ESC-derived neurons compared to the isogenic control cell line (Mann-Whitney U test = 6, p = 0.6857). (**C**) No differences in pTau-T181 levels were observed in ESC-derived cortical neurons between HD and control cell lines (Mann-Whitney U test = 7, p = 0.8857). (**D**) Representative western blot images of pTau-S404, pTau-T181, and GAPDH in HD and control NSCs demonstrating no differences in pTau-S404 and pTau-T181 levels. (**E**) There were no changes in pTau-S404 levels in HD NSCs compared to isogenic control NSCs (Mann-Whitney U test = 4, p > 0.9999). (**F**) pTau-T181 levels were not changed in HD NSCs compared to isogenic control cells (Mann-Whitney U test = 2, p = 0.4000).

**Supplementary Figure 6.**
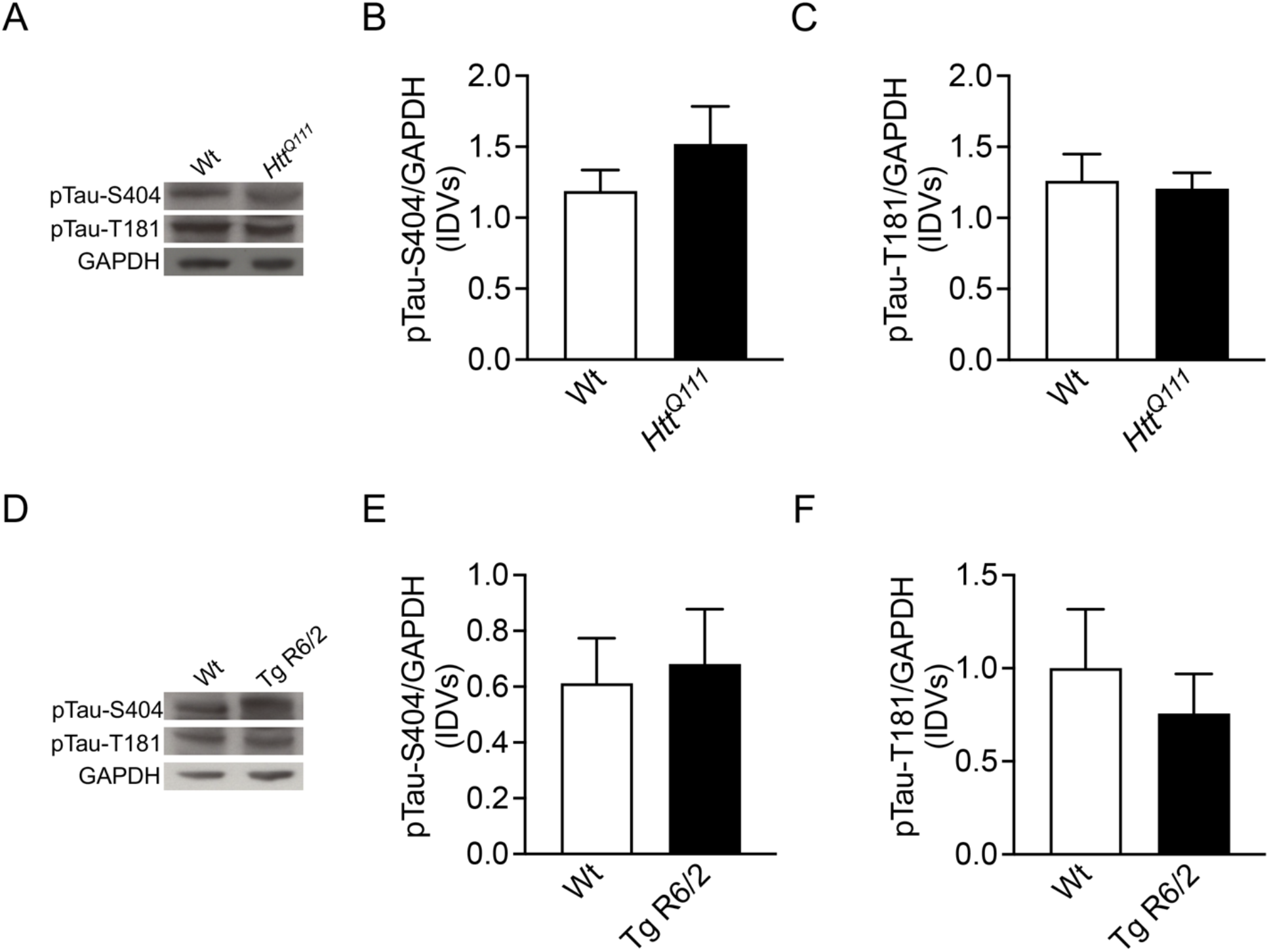
There were no alterations in pTau-S404 and pTau-T181 levels in two HD mouse models. (**A**) Representative western blot images of pTau-S404, pTau-T181 and GAPDH in wild-type (Wt) and *Htt^Q111^* cortex revealing no alterations in pTau-S404 and pTau-T181 levels. (**B**) There was no change in pTau-S404 levels in *Htt^Q111^* (n = 8) compared to Wt cortex (n = 8) (Mann-Whitney U test = 26, p = 0.5737) (**C**) There were no alterations in pTau-T181 levels in *Htt^Q111^* (n = 8) compared to Wt cortex (n = 8) (Mann-Whitney U test = 32, p > 0.9999). (**D**) Representative western blot images of pTau-S404, pTau-T181 and GAPDH in Tg and Wt R6/2 mice demonstrating no differences in pTau-S404 and pTau-T181 levels. (**E**) No alterations in pTau-S404 levels were observed in Tg R6/2 (n = 8) compared to Wt cortex (n = 8) (Mann-Whitney U test = 25, p = 0.77789). (**F**) There were no alterations in pTau-T181 levels in Tg R6/2 (n = 8) compared to Wt cortex (n = 8) (Mann-Whitney U test = 32, p > 0.9999).

